# Plasma GFAP associates with secondary Alzheimer’s pathology in Lewy body disease

**DOI:** 10.1101/2022.12.05.22283106

**Authors:** Katheryn A.Q. Cousins, David J. Irwin, Alice Chen-Plotkin, Leslie M. Shaw, Sanaz Arezoumandan, Edward B. Lee, David A. Wolk, Daniel Weintraub, Meredith Spindler, Andres Deik, Murray Grossman, Thomas F. Tropea

**Author notes:** Correspondence should be addressed to: Katheryn Cousins, Penn Frontotemporal Degeneration Center, Department of Neurology, Richards Medical Research Laboratories, Suite 600B, 3700 Hamilton Walk, Philadelphia, PA 19104.

## Abstract

**Objective:** Within Lewy body spectrum disorders (LBSD) with α-synuclein pathology (αSyn), concomitant Alzheimer’s disease (AD) pathology is common and is predictive of clinical outcomes, including cognitive impairment and decline. Plasma phosphorylated tau 181 (p-tau_181_) is sensitive to AD neuropathologic change (ADNC) in clinical AD, and plasma glial fibrillary acidic protein (GFAP) is associated with the presence of β-amyloid plaques. While these plasma biomarkers are well tested in clinical and pathological AD, their diagnostic and prognostic performance for concomitant AD in LBSD is unknown.

**Methods:** In autopsy-confirmed αSyn-positive LBSD, we tested how plasma p-tau_181_ and GFAP differed across αSyn with concomitant ADNC (αSyn+AD; n=19) and αSyn without AD (αSyn; n=30). Severity of burden was scored on a semi-quantitative scale for several pathologies (*e.g*., β-amyloid and tau), and scores were averaged across sampled brainstem, limbic, and neocortical regions.

**Results:** Linear models showed that plasma GFAP was significantly higher in αSyn+AD compared to αSyn (β=0.31, 95%CI=0.065 – 0.56, *p*=0.015), after covarying for age at plasma, plasma-to-death interval and sex; plasma p-tau_181_ was not (*p*=0.37). Next, linear models tested associations of AD pathological features with both plasma analytes, covarying for plasma-to-death, age at plasma, and sex. GFAP was significantly associated with brain β-amyloid (β=15, 95%CI=6.1 – 25, *p*=0.0018) and tau burden (β=12, 95%CI=2.5 – 22, *p*=0.015); plasma p-tau_181_ was not associated with either (both *p*>0.34).

**Interpretation:** Findings indicate that plasma GFAP may be sensitive to concomitant AD pathology in LBSD, especially accumulation of β-amyloid plaques.

## 1. Introduction

Lewy body spectrum disorders (LBSD) are a group of movement disorders that include Parkinson’s disease (PD), PD with dementia (PDD), and dementia with Lewy bodies (DLB). While α-synuclein (αSyn) is the primary pathology associated with LBSD, concomitant Alzheimer’s disease (AD) pathology is common. Nearly 50% of cases have significant accumulations of β-amyloid plaques and tau neurofibrillary tangles (intermediate or high AD neuropathologic change [ADNC]), justifying a secondary diagnosis of AD at autopsy^1,2^. Concomitant AD may describe much of the clinical heterogenity across LBSD^3^, and is associated with memory and linguistic impairments^4–6^, postural instability and gait^7^, and reduced survival^2,6^. Thus, the *in vivo* detection of concomitant AD is important for prognosis and disease management of LBSD^8^ and may inform clinical trial design^9^. Yet, biofluid markers that are well-established in canonical AD must still be validated in LBSD to test sensitivity to concomitant ADNC. Two factors may affect biomarker accuracy to detect secondary AD. First, overall burden of ADNC is relatively less severe in LBSD compared to AD, with high proportions of intermediate ADNC^1,2^. Second, cerebrospinal fluid (CSF) biomarkers of AD have an altered profile in LBSD. In particular, CSF tau phosphorylated at threonine 181 (p-tau_181_) was inversely associated with αSyn pathology, which reduced its sensitivity to concomitant AD^10^. Another study showed that CSF p-tau_181_ was significantly lower in early PD than controls, further evincing that concentrations are reduced in LBSD^11^. Thus, as plasma biomarkers for AD are developed, it becomes important to validate their efficacy to detect AD co-pathology in LBSD.

Two candidate AD plasma biomarkers in LBSD are plasma p-tau_181_ and glial fibrillary acidic protein (GFAP). Several studies in autopsy and living participants have shown that plasma p-tau_181_ can detect AD^12,13^ and is associated with accumulation and spread of brain β-amyloid and tau pathology^13^. GFAP is a cytoskeletal filament protein highly expressed in astrocytes, and concentrations of GFAP are raised in the CSF and blood following astrogliosis and the degeneration of astrocytes^14^. Neuroinflammation plays a key role in AD pathogenesis^15^, and plasma GFAP may be an early marker correlated with brain β-amyloid pathology^16,17^. Still, the majority of work has studied both analytes in the context of primary AD. Utility of these plasma biomarkers in LBSD is unclear, due to different findings across studies of living LBSD, which define groups using clinical or positron emission tomography (PET) data^18–20^. In light of these conflicting findings in biomarker-defined LBSD, it becomes necessary to test plasma biomarkers in autopsy cases with pathologically-confirmed diagnoses.

In this autopsy study, we compare plasma p-tau_181_ and GFAP in αSyn with concomitant AD (αSyn+AD) *vs*. αSyn without AD; AD without concomitant αSyn is included as a reference group. Multivariable models test how each analyte correlates with brain accumulation of pathological β-amyloid plaques and tau neurofibrillary tangles, as well as αSyn deposition and gliosis. Receiver operating characteristic (ROC) analyses test how accurately analytes detect ADNC in this mixed pathology sample, and we also tested associations with global cognition (mini mental state exam [MMSE]).

## 2. Methods

### 2.1 General Selection Criteria

Participants were enrolled at the University of Pennsylvania (Penn) Parkinson’s Disease Research Center, Frontotemporal Degeneration Center, or Alzheimer’s Disease Research Center, and were selected retrospectively from the Penn Integrated Neurodegenerative Disease Database^21^ on September 15, 2022, based on eligibility criteria outlined below. Written consent was obtained, and approved by the Penn Institutional Review Board.

Selection criteria were clinically diagnosed LBSD with neuropathological diagnoses of either αSyn (n=30) or αSyn+AD (n=19), and available biomarkers of either plasma GFAP or plasma p-tau_181_. As a neuropathological reference group, we also examined autopsy-confirmed AD without αSyn (n=21), with a clinical diagnosis of AD, and available plasma GFAP and p-tau_181_. If individuals had plasma collected at 2 or more timepoints, the last timepoint was selected to more closely reflect pathology at autopsy. There was a median interval of 2 years (interquartile range [IQR]=2; max=11) between plasma collection and death. Finally, we used propensity score matching^22^ to select 70 clinically normal individuals as Controls, matched for age and sex, with an MMSE≥27; Controls did not have autopsy data.

### 2.2 Neuropathological diagnoses and assessments

Brains were sampled at autopsy and assessed for ADNC and αSyn, as well as for other pathologies according to standardized procedures^21,23^. ADNC was scored according to ABC criteria^24^ and high or intermediate ADNC was considered AD positive; low or not ADNC was considered AD negative. Thal phase^25^, Braak stage^26^, and CERAD score^27^ are reported using a 4-point scale (0-3)^24^. DLB stage was assessed^28^ and brainstem predominant, limbic, and neocortical Lewy bodies were all considered αSyn positive; no or amygdala predominant Lewy bodies were considered αSyn negative. In addition to ADNC and αSyn, the presence of concomitant vascular disease^29^ and TDP-43^30^ was assessed; only 3 LBSD patients total had moderate or high vascular disease (1 αSyn+AD; 2 αSyn)^29^ and it was therefore not assessed in analyses.

Brain tissue samples were stained using immunohistochemistry as previously described^21^, and gross severity of pathological accumulations of β-amyloid, tau, αSyn, and TDP-43 were scored using a semi-quantitative scale (0=none, 0.5=rare, 1=minimal, 2=moderate, 3=severe); in addition severity of gliosis and cerebral amyloid angiopathy (CAA) were likewise quantified. Burden scores were the average across regions standardly sampled^24^, which included the amygdala, cingulate, CA1/subiculum, endorhinal cortex, middle frontal gyrus, angular gyrus, superior/middle temporal gyrus, pons, and medulla. Hemisphere was randomized; if both hemispheres were sampled, the average was taken. Missing regional data were dropped to calculate the average.

### 2.3 Plasma analysis

Plasma was collected and assayed for p-tau_181_ and GFAP previously described^31,32^. Plasma samples were analyzed on the Quanterix HD-X automated immunoassay platform. Samples were analyzed in duplicate using the Discovery kit reagents for GFAP^33^ and using the V2 Advantage kit for p-tau_181_^31^.

In the autopsy sample, 1 αSyn and 1 AD were missing plasma GFAP.

### 2.4 Clinical and demographic features

Demographic features were available through INDD. Where applicable, we examined age at onset (first reported symptom; years), age at plasma collection (years), disease duration at plasma (time from onset to plasma collection; years), interval from plasma-to-CSF (years), interval from plasma-to-MMSE (years), interval from plasma-to-death (years), and age at death (years). Sex and race were determined by self-report.

MMSE was used as a measure of global cognition in LBSD that was available in historical autopsy cases. In the LBSD autopsy sample, 24 αSyn and 16 αSyn+AD patients had MMSE available. There was a median interval of 0.6 years (IQR=2; max=8.2) between plasma collection and MMSE.

### 2.5 Statistical Analyses

Not all demographic and analyte variables were normally distributed, therefore non-parametric Kruskal-Wallis and Mann-Whitney-Wilcoxon tests performed uncorrected group comparisons for continuous variables. Chi-square tests performed group comparisons for categorical variables. Spearman correlations tested within-group associations between continuous variables, with nominal and Bonferroni-corrected *p*-values reported. All statistical models used a significance threshold of α=0.05.

For linear models, 95% confidence intervals (CI) for β-estimates were reported. Plasma p-tau_181_ and GFAP were not normally distributed and were log-transformed in all models. Distributions of pathological and clinical measures varied, and were therefore rank-transformed to perform non-parametric analyses. Effect sizes with 95%CI were calculated using generalized η^2^ (η^2^_*G*_)^34^, using standard interpretation cutoffs (≥0.01 small, ≥0.06 medium, ≥0.14 large)^35^. Statistical analyses were performed and figures were generated using R version 4.1.2 (2021-11-01).

First, linear models tested how plasma biomarkers differed across αSyn and αSyn+AD, covarying for age at plasma collection, interval from plasma-to-death, and sex (Equation 1).

1. *log*(Plasma Analyte) = β_0_ + β_1_*Group + β_2_*Age + β_3_*Plasma-to-Death + β_4_*Sex + ε Within LBSD, linear models tested how AD pathological hallmarks β-amyloid plaques (Equation 2) and tau neurofibrillary tangles (Equation 3), associated with plasma GFAP and plasma p-tau_181_, covarying for age, interval from plasma-to-death, and sex. Models were repeated excluding individuals with a plasma-to-death interval > 5 years.
2. *rank*(β-amyloid) = β_0_ + β_1_\**log*(p-tau_181_) + β_2_\**log*(GFAP) + β_3_*Age + β_4_*Plasma-to-Death + β_5_*Sex + ε
3. *rank*(Tau) = β_0_ + β_1_\**log*(p-tau_181_) + β_2_\**log*(GFAP) + β_3_*Age + β_4_*Plasma-to-Death + β_5_*Sex + ε Given correlations between β-amyloid, tau and gliosis burden in LBSD, a post-hoc model tested associations of all with plasma GFAP (Equation 4), covarying for for age, interval from plasma-to-death, and sex.
4. *log*(GFAP) = β_0_ + β_1_\**rank*(β-amyloid) + β_2_\**rank*(Tau) + β_3_\**rank*(Gliosis) + β_4_*Age + β_5_*Plasma-to-Death + β_6_*Sex + ε ROC analyses using bootstrapping (500 iterations) compared discrimination of High/Intermediate ADNC from Not/Low ADNC; AUC with 90%CIs were reported^36^. ROC analyses tested plasma analytes in the full autopsy sample (αSyn, αSyn+AD, and AD) and within LBSD (αSyn and αSyn+AD). Finally, we tested associations of global cognition (MMSE, rank-transformed) with both analytes, covarying for age, MMSE-to-plasma interval (years), and sex. This analysis was repeated excluding individuals with a plasma-to-MMSE interval > 1 year.
5. *rank*(MMSE) = β_0_ + β_1_\**log*(GFAP) + β_2_\**log*(p-tau_181_) + β_3_*Age + β_4_*MMSE-to-plasma + β_5_*Sex + ε

## 3. Results

Table 1 summarizes group characteristics and Kruskal-Wallis group-wise comparisons. Pairwise Wilcoxon tests show that αSyn+AD had an older median age of onset than αSyn (W=137.5, *p*=0.0025); there was no difference in age at plasma (*p*=0.37), plasma-to-death interval (*p*=0.76), or age at death (*p*=0.37). Chi-square tests showed no difference in sex (*p*=0.32). Compared to AD, αSyn+AD had lower Thal Phase (χ^2^=5, *p*=0.039), Braak Stage (χ^2^=18, *p*=0.00050), and CERAD score (χ^2^=9.1, *p*=0.010). The proportion of APOE ε4 alleles did not differ between AD and αSyn+AD (*p*=0.42).

**Table 1:** Demographic, pathological, and clinical characteristics of participants. For continuous variables, median and interquartile range (IQR) are reported; Kruskal-Wallis tests performed group comparisons. For categorical variables, count (percentage [%]) are provided; chi-square tests performed frequency comparisons. *p*-values are reported for group comparisons. Note that DLB Type and ADNC (including Thal, Braak, and CERAD) were used to define groups.

| | Control | αSyn | αSyn+AD | AD | p |
| --- | --- | --- | --- | --- | --- |
| n | 70 | 30 | 19 | 21 |  |
| Age at Onset (years) | -- | 60 [54, 66] | 69 [62, 74] | 62 [59, 66] | 0.009 |
| Age at Plasma (years) | 72 [65, 77] | 75 [68, 79] | 74 [70, 84] | 71 [64, 75] | 0.083 |
| Duration at Plasma (years) | -- | 13 [9, 16] | 7 [4, 10] | 5 [4, 8] | <0.001 |
| Plasma to Death (years) | 4 [2, 5] | 2 [1, 2] | 2 [1, 2] | 3 [2, 4] | 0.095 |
| Age at Death (years) | 73 [70, 79] | 77 [70, 81] | 76 [72, 84] | 73 [69, 77] | 0.233 |
| Sex = Male (%) | 39 (56%) | 21 (70%) | 16 (84%) | 11 (52%) | 0.079 |
| Self-reported Race (%) |  |  |  |  | 0.025 |
| Asian | 2 (3%) | 0 (0%) | 0 (0%) | 0 (0%) |  |
| Black or African American | 16 (23%) | 0 (0%) | 0 (0%) | 2 (10%) |  |
| More than One Race | 1 (1%) | 0 (0%) | 0 (0%) | 1 (5%) |  |
| White | 50 (72%) | 30 (100%) | 19 (100%) | 18 (86%) |  |
| Thal Phase (0-3) | -- | 1 [0, 1] | 3 [2, 3] | 3 [3, 3] | <0.001 |
| Braak Stage (0-3) | -- | 1 [1, 2] | 2 [2, 3] | 3 [3, 3] | <0.001 |
| CERAD Score (0-3) | -- | 0 [0, 1] | 2 [2, 3] | 3 [3, 3] | <0.001 |
| ADNC (%) |  |  |  |  | <0.001 |
| Not | -- | 11 (37%) | 0 (0%) | 0 (0%) |  |
| Low | -- | 19 (63%) | 0 (0%) | 0 (0%) |  |
| Intermediate | -- | 0 (0%) | 14 (74%) | 1 (5%) |  |
| High | -- | 0 (0%) | 5 (26%) | 20 (95%) |  |
| DLBType (%) |  |  |  |  | <0.001 |
| None | -- | 0 (0%) | 0 (0%) | 10 (48%) |  |
| Amygdala Predominant | -- | 0 (0%) | 0 (0%) | 11 (52%) |  |
| Brainstem Predominant | -- | 6 (20%) | 1 (5%) | 0 (0%) |  |
| Transitional or Limbic | -- | 15 (50%) | 5 (26%) | 0 (0%) |  |
| Diffuse or Neocortical | -- | 9 (30%) | 13 (68%) | 0 (0%) |  |
| Vascular disease+ (%) | -- | 2 (7%) | 1 (5%) | 3 (14%) | 0.527 |
| APOE ε4 (%) | | | | | <0.001 |
| 0 | 49 (72%) | 19 (63%) | 6 (33%) | 6 (29%) |  |
| 1 | 17 (25%) | 11 (37%) | 10 (56%) | 9 (43%) |  |
| 2 | 2 (3%) | 0 (0%) | 2 (11%) | 6 (29%) |  |
| Clinical Diagnosis (%) |  |  |  |  | <0.001 |
| Control | 70 (100%) | 0 (0%) | 0 (0%) | 0 (0%) |  |
| AD | 0 (0%) | 0 (0%) | 0 (0%) | 18 (95%) |  |
| PD/PDD | 0 (0%) | 27 (90%) | 11 (58%) | 0 (0%) |  |
| DLB | 0 (0%) | 3 (10%) | 8 (42%) | 1 (5%) |  |

### 3.1 Group comparisons

To test which plasma analytes associate with concomitant AD pathology, Figure 1 compares analyte levels across αSyn and αSyn+AD, with AD as a reference group. Linear models covarying for age at plasma, plasma-to-death interval, and sex confirmed that GFAP was significantly higher in αSyn+AD than αSyn (β=0.31, 95%CI=0.065 – 0.56, *p*=0.015) with large effect size (η^2^_*G*_=0.14; 95%CI=0.021 – 1.0); plasma p-tau_181_ was not significantly higher in αSyn+AD than αSyn (*p*=0.37).

**Figure 1.**
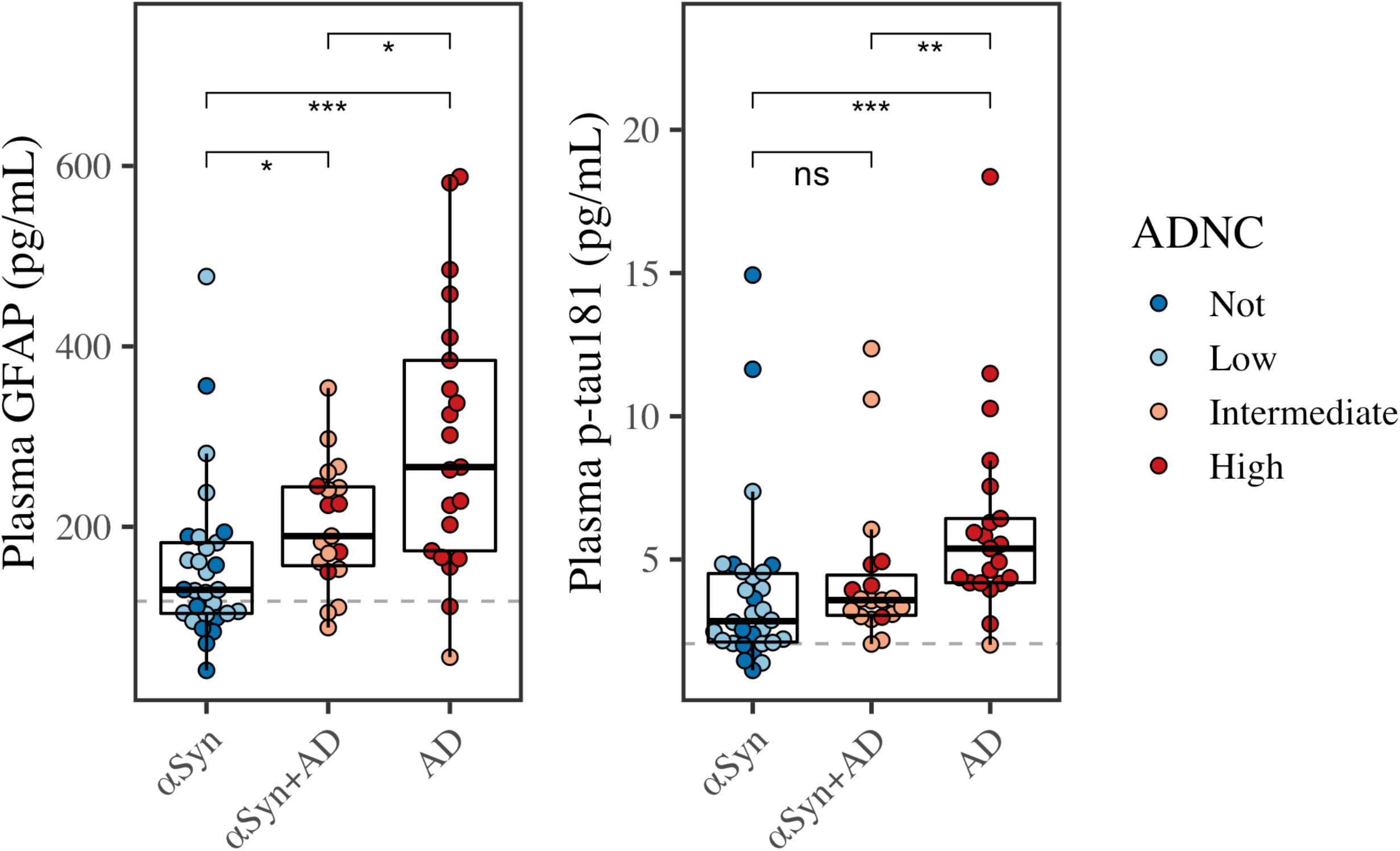
Plasma concentrations across pathological diagnosis. Boxplots show median, interquartile range (IQR), and outliers for plasma p-tau_181_, GFAP, and NfL. Color represents ADNC. Horizontal dashed lines plot medians for GFAP and p-tau_181_ from neurologically normal Controls. Asterisks represent *p*-values from Wilcoxon pairwise comparisons (* *p*<0.05, ** *p*<0.01, *** *p*<0.001, and not significant [ns]).

Examining covariates, age at plasma was associated with GFAP (β=0.017, 95%CI=0.00094 – 0.034, *p*=0.039) but not p-tau_181_ (*p*=0.27). Plasma-to-death was associated with GFAP (β=-0.056, 95%CI=-0.10 – -0.0078, *p*=0.024) but not p-tau_181_ (*p*=0.61). Sex was associated with neither GFAP (*p*=0.16) nor p-tau_181_ (*p*=0.12).

Compared to neurologically healthy Controls, Wilcoxon tests showed that plasma GFAP was higher in αSyn+AD (W=229, *p*=0.000013) and AD groups (W=174, *p*=1.3e-07), but not αSyn (W=761.5, *p*=0.052). Compared to Controls, plasma p-tau_181_ was higher in all three groups: αSyn+AD (W=230.5, *p*=0.000014), AD (W=125, *p*=9.4e-09), and αSyn (W=623, *p*=0.0013).

In the full autopsy sample (αSyn, αSyn+AD, AD), Figure 2 tests plasma analytes across metrics of AD severity: ADNC, Thal phase, and Braak stage.

**Figure 2.**
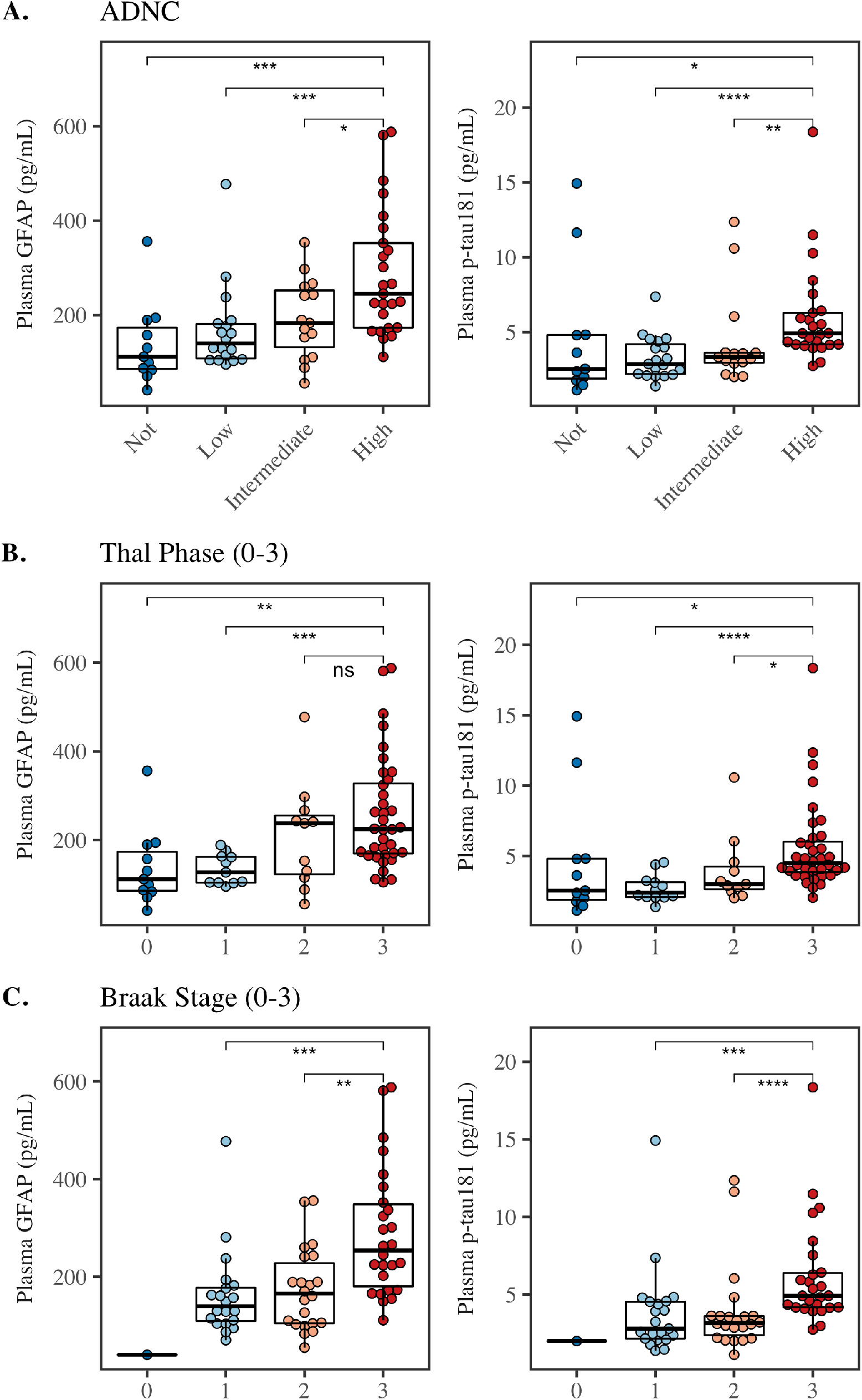
Plasma concentrations across ADNC, Thal Phase, and Braak Stage across all autopsy cases (αSyn, αSyn+AD, AD). Plasma GFAP (left panels) and p-tau_181_ (right panels) comparisons across (A.) ADNC score, (B.) Thal Phase, and (C.) Braak stage. Boxplots show median, interquartile range (IQR), and outliers. Color represents severity. Asterisks represent *p*-values from Wilcoxon pairwise comparisons (* *p*<0.05, ** *p*<0.01, *** *p*<0.001, **** *p*<0.0001, and not significant [ns]).

### 3.2 Associations with pathological accumulation within LBSD

Within LBSD, we explored the pathological correlates (β-amyloid, tau, gliosis, and αSyn) of plasma GFAP and p-tau_181_ (Figure 3); we also tested associations with TDP-43 and CAA to ensure other pathologies were not influencing plasma concentrations.

**Figure 3.**
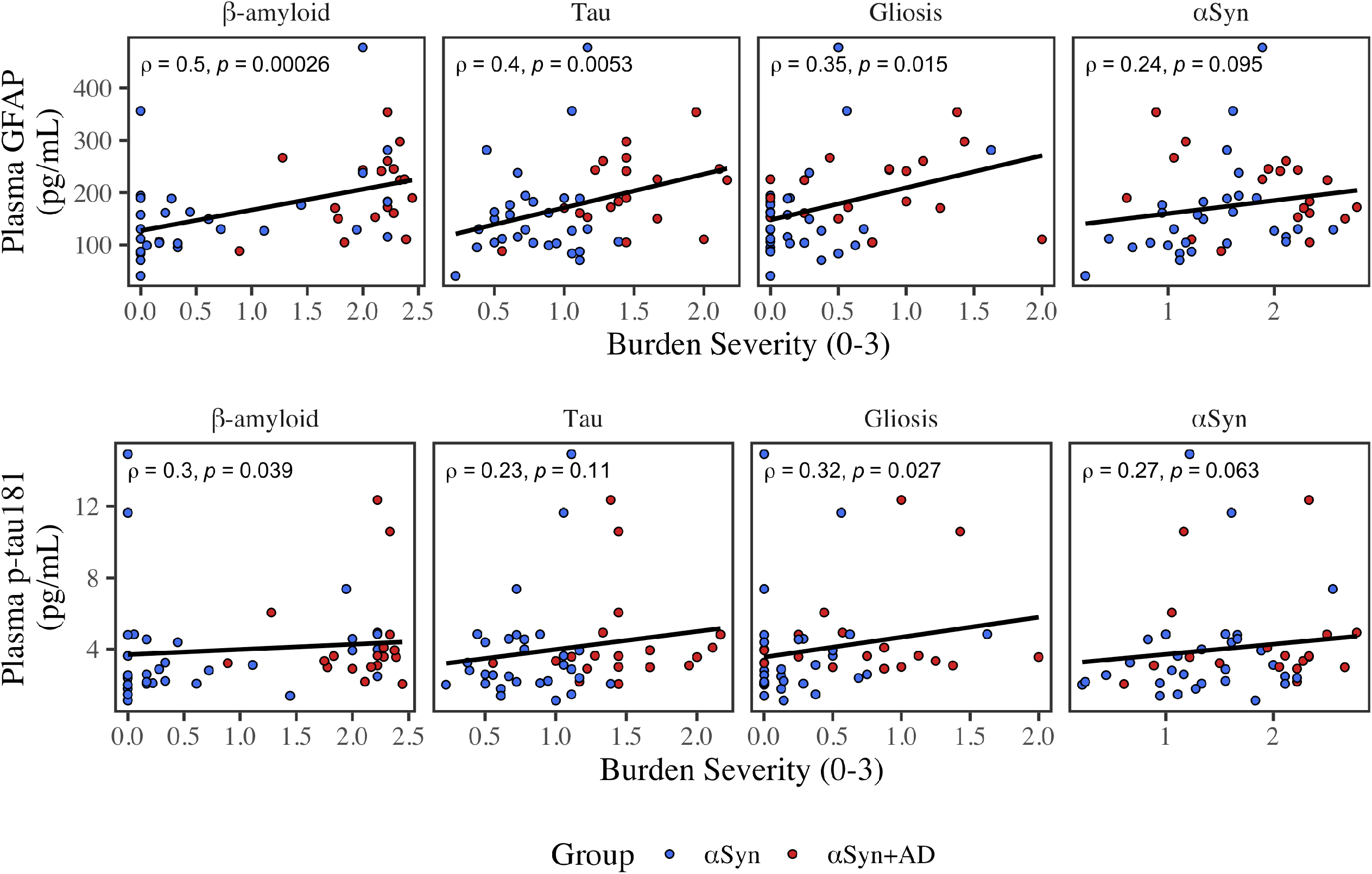
Correlations of plasma analytes and measures of pathological burden. Scatterplots for plasma GFAP (top panel) and p-tau_181_ (bottom panel) and pathological variables are plotted. Color represents αSyn (blue) and αSyn+AD (red). Least squares regression lines are plotted in black. Spearman’s rho (ρ) and nominal *p*-values are reported.

Across all LBSD, plasma GFAP was positively associated with β-amyloid (rho=0.5, *p*=0.00026), tau (rho=0.4, *p*=0.0053), and gliosis (rho=0.35, *p*=0.015); plasma GFAP was not associated with αSyn (rho=0.24, *p*=0.095), TDP-43 (*p*=0.92), or CAA (*p*=0.78). Association of plasma GFAP with β-amyloid survived multiple corrections (*p*=0.0058), but not tau (*p*=0.071) or gliosis (*p*=0.062).

Plasma p-tau_181_ was positively associated with β-amyloid (rho=0.30, *p*=0.039) and gliosis (rho=0.32, *p*=0.027); plasma p-tau_181_ was not associated with tau (*p*=0.11), αSyn (rho=0.27, *p*=0.063), TDP-43 (*p*=0.24), or CAA (rho=0.25, *p*=0.079). However, associations with β-amyloid (*p*=1.0) and gliosis (*p*=1.0) did not survive multiple correction.

In addition, plasma GFAP and p-tau_181_ were correlated with each other within LBSD (rho=0.31, *p*=0.031).

Next we tested how both analytes associated with AD pathological hallmarks within LBSD. A linear model tested β-amyloid plaque as a function of both plasma p-tau_181_ and GFAP, covarying for age at plasma collection, plasma-to-death interval, and sex. β-amyloid plaque burden was significantly associated with plasma GFAP (β=15, 95%CI=6.1 – 25, *p*=0.0018) with large effect size (η^2^_*G*_=0.23; 95%CI=0.066 – 1.0), but not plasma p-tau_181_ (*p*=0.95). Results were robust after excluding individuals who had plasma collection > 5 years before death (GFAP: β=14, 95%CI=3.7 – 24, *p*=0.0089; p-tau_181_: *p*=0.82).

Likewise, we tested pathological tau burden as a function of both plasma p-tau_181_ and GFAP, covarying for age at plasma collection, plasma-to-death interval, and sex. Pathological tau burden was significantly associated with plasma GFAP (β=12, 95%CI=2.5 – 22, *p*=0.015) with large effect size (η^2^ =0.23; 95%CI=0.066 – 1), but not plasma p-tau_181_ (*p*=0.34). Results were robust after excluding individuals who had plasma collection > 5 years before death (GFAP: β=12, 95%CI=2.1 – 23, *p*=0.020; p-tau_181_: *p*=0.30).

β-amyloid plaque burden was not significantly associated with any of the covariates: age (*p*=0.99), plasma-to-death (*p*=0.62), or sex (*p*=0.10). Tau burden was not associated with age (*p*=0.30), plasma-to-death (β=1.5, 95%CI=-0.075 – 3.1, *p*=0.061), or sex (*p*=0.81).

Pathological accumulations of β-amyloid, tau, and gliosis were all positively correlated in LBSD (β-amyloid & tau: rho=0.51, *p*=0.00017; β-amyloid & gliosis: rho=0.32, *p*=0.025; tau & gliosis: rho=0.46, *p*=0.001). Given that plasma GFAP was positively associated with β-amyloid, tau, and gliosis burden (Figure 3) and thus the potential for collinearity, a post-hoc linear model tested for plasma GFAP as a function of all three pathologies (β-amyloid, tau, and gliosis burden) to determine which might independently associate with plasma GFAP levels. In addition, age at plasma, plasma-to-death interval, and sex were included as covariates. Plasma GFAP was significantly associated with β-amyloid burden (β=0.011, 95%CI=0.0011 – 0.02, *p*=0.030) with large effect size (η^2^_*G*_=0.3; 95%CI=0.12 – 1.0); however, GFAP was not associated with gliosis (*p*=0.35) or tau (*p*=0.41) after covarying for β-amyloid. In this model, covariates age (β=0.014, 95%CI=-0.0021 – 0.030, *p*=0.086), plasma-to-death (β=-0.043, 95%CI=-0.090 – 0.0041, *p*=0.073), and sex (*p*=0.15) were not significant.

### 3.3 ROC analyses

ROC analyses (Figure 3) tested the diagnostic accuracy of both analytes to detect ADNC (high/intermediate) in the full sample (αSyn, αSyn+AD, AD) and again in LBSD αSyn positive cases (αSyn, αSyn+AD). Neither analyte demonstrated high diagnostic performance in LBSD: for the full sample, plasma GFAP had an AUC of 0.77 (90%CI=0.67 – 0.86) and plasma p-tau_181_ had an AUC of 0.72 (90%CI=0.61 – 0.82). In the LBSD sample, plasma GFAP had an AUC of 0.71 (90%CI=0.58 – 0.83) and plasma p-tau_181_ had an AUC of 0.64 (90%CI=0.50 – 0.77).

### 3.4 Clinical correlation with MMSE

In LBSD, we tested how plasma analytes associated with cognition; Spearman’s correlations show that both plasma GFAP and p-tau_181_ were correlated with MMSE (Figure 5). In a model covarying for age, plasma-to-MMSE interval and sex, lower MMSE was associated with higher GFAP (β=-8.2, 95%CI=-15 – -1.2, *p*=0.024), but not p-tau_181_ (*p*=0.55). We repeated the model, excluding cases with plasma-to-MMSE interval > 1 year: results were consistent with lower MMSE significantly associated with higher GFAP (β=-12, 95%CI=-21 – -1.8, *p*=0.023), but not p-tau_181_ (*p*=0.44).

**Figure 4.**
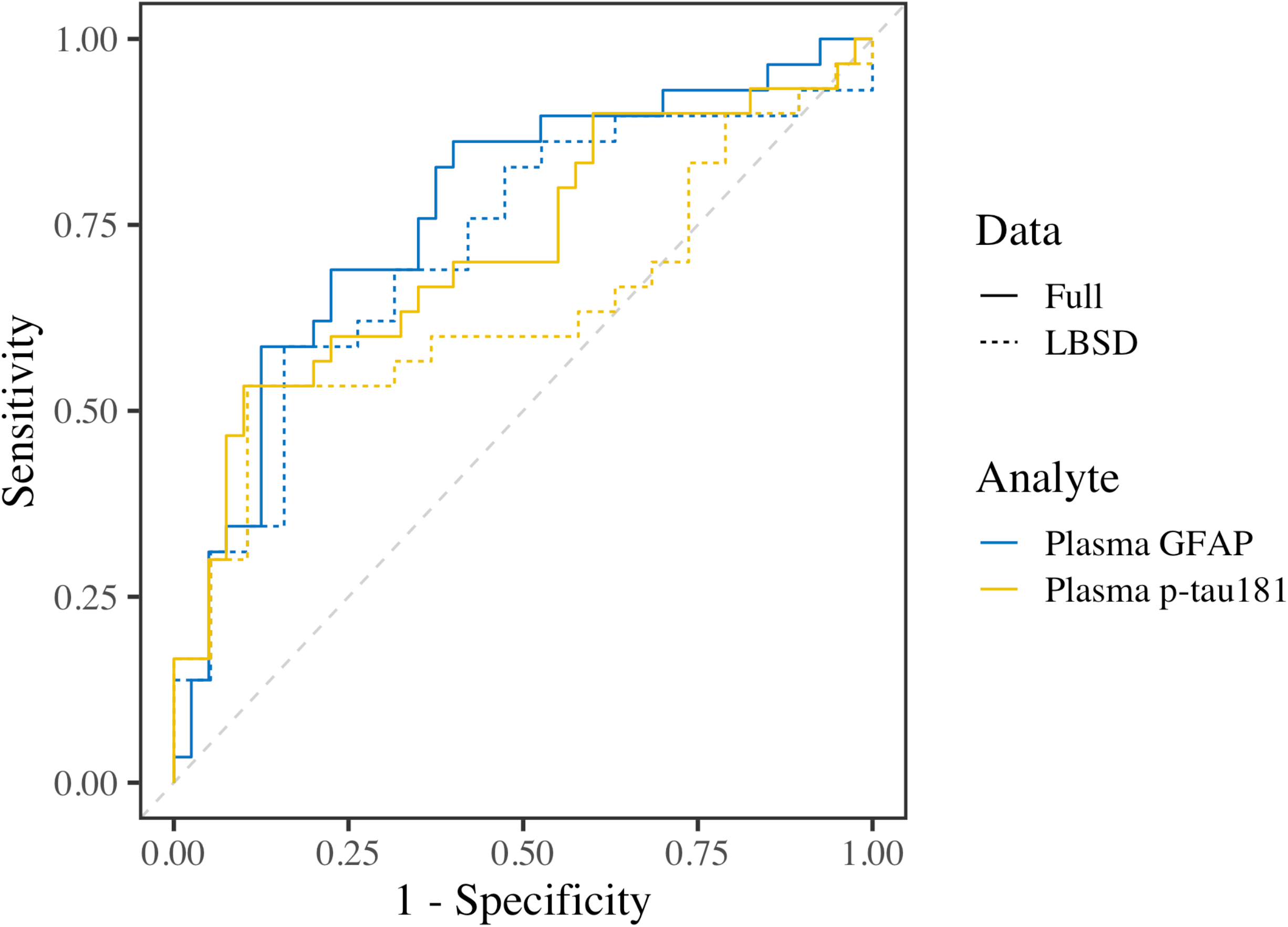
Receiver operating characteristic (ROC) curves. Color indicates plasma GFAP (blue) and p-tau_181_ (yellow). Solid lines represent the full autopsy dataset (αSyn, αSyn+AD, AD), and dotted lines represent LBSD αSyn positive cases (αSyn, αSyn+AD). The dashed grey line represents chance performance. AUCs were 0.77 for full-sample GFAP, 0.71 for LBSD-sample GFAP, 0.72 for full-sample p-tau_181_, and 0.64 for LBSD-sample p-tau_181_.

**Figure 5.**
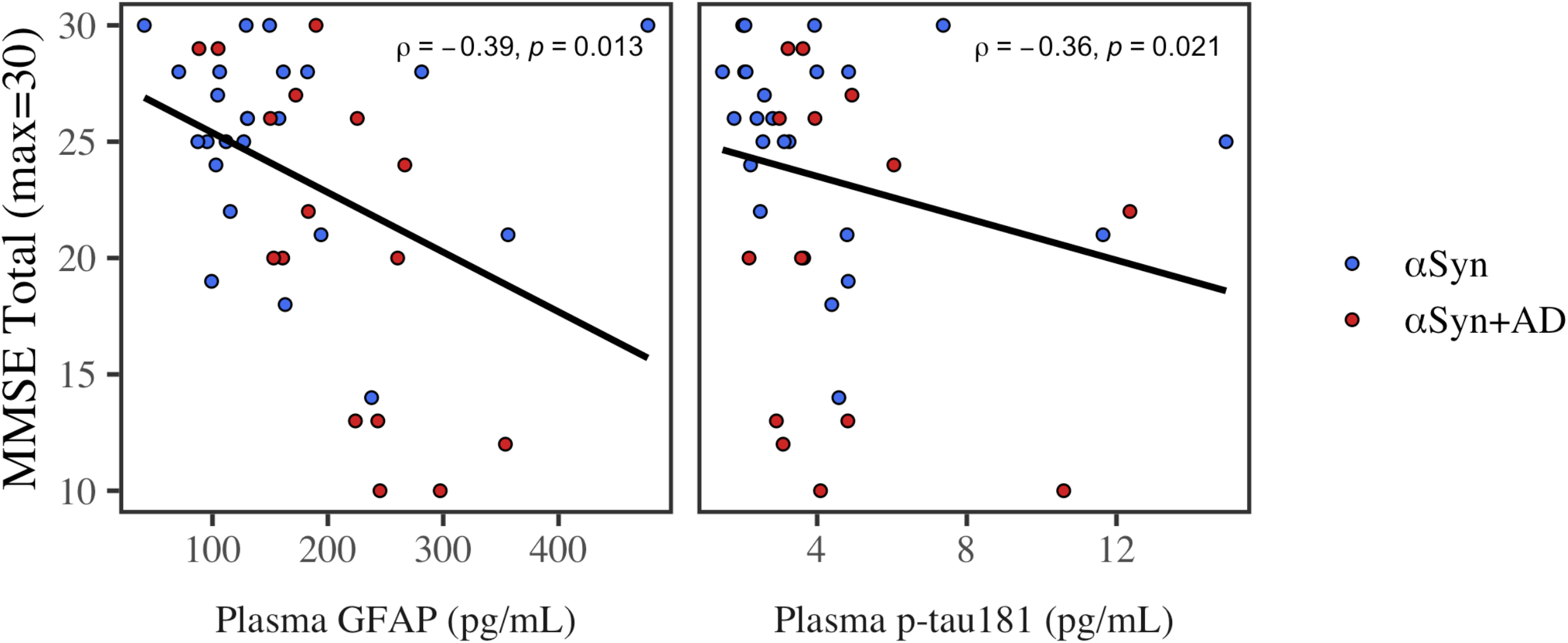
Associations between analytes and MMSE. Color indicates LBSD group αSyn (blue) and αSyn+AD (red). Least squares regression line is plotted. Spearman’s rho (ρ) and *p*-value are reported.

Examining covariates, MMSE was associated with plasma to MMSE interval (β=-2.1, 95%CI=-3.2 – -0.89, *p*=0.001) and sex (β=-7.8, 95%CI=-15 – -0.14, *p*=0.046), but was not associated with age (*p*=0.70).

## Discussion

Accumulating evidence shows that plasma biomarkers, such as p-tau_181_ and GFAP, are highly sensitive to AD^12,13,37^. Yet, plasma biomarkers have not been well examined in the context of LBSD with concomitant ADNC. In this autopsy study, we show consistent results that plasma GFAP is sensitive to concomitant ADNC in LBSD with autopsy-confirmed αSyn: antemortem plasma GFAP was significantly higher in αSyn+AD than αSyn, it was associated with higher burden of postmortem β-amyloid (even after covarying for gliosis), and was associated with worse antemortem MMSE performance. Surprisingly, we do not find as robust results for plasma p-tau_181_. Plasma p-tau_181_ was not higher in αSyn+AD than αSyn. Postmortem pathological associations with β-amyloid plaques and neurofibrillary tau showed a stronger association with plasma GFAP than p-tau_181_. Our findings emphasize conclusions from previous studies that LBSD specific strategies may be necessary to detect concomitant AD^8,10,38^, and that plasma biomarkers show unique profiles that may diverge from those seen in CSF^39,40^. Our results here suggest that plasma GFAP is a promising biomarker that is sensitive to concomitant AD pathology in LBSD, and may reflect accumulation of β-amyloid pathology.

We note that ADNC is typically less severe in LBSD cases than clinical AD^1,2^. In this study, 74% of αSyn+AD in this sample were intermediate ADNC (26% were high ADNC). In AD a higher percentage were high ADNC (95%), and likewise AD cases showed significantly higher plasma p-tau_181_ than αSyn+AD. When examining ADNC, Thal phase, and Braak stage, plasma p-tau_181_ appeared elevated only at the highest levels (*i.e*., high ADNC, Thal/Braak=3); our findings echo other research showing plasma p-tau_181_ is elevated only at more severe AD stages^41^, and is significantly lower in intermediate ADNC than high ADNC cases^42^. Likewise, studies that demonstrate good diagnostic accuracy of plasma p-tau_181_ in autopsy samples have typically tested discrimination of high ADNC from intermediate/low/not ADNC, showing ROC AUCs of 0.77 – 0.91^12,13,43^. Longitudinal studies show that this relationship may depend on time between plasma collection and death^37^. An open question is whether other epitopes of p-tau (such as 217 or 231) are more sensitive to intermediate ADNC, especially at earlier AD pathophysiological stages^13,41,42,44^. Poor sensitivity of plasma p-tau_181_ to intermediate ADNC may help explain the divergences between previous findings: one study showed no difference in plasma p-tau_181_ in LBSD who were PET-Aβ positive versus PET-Aβ negative^18^, although others have found that plasma p-tau_181_ does associate with PET-tau status in LBSD^19^ and that it associates with cognitive decline in DLB^20^. Another possible factor is that biomarkers, like CSF and PET, can show altered profiles in LBSD compared to AD^10,38^. There is some rare autopsy work examining effects of concomitant pathology on plasma biomarkers in primary AD^37,42^; findings demonstrate significantly higher plasma p-tau_181_ in primary AD patients with mixed pathology, including concomitant Lewy bodies, than non-AD. Future studies will be needed to disentangle these contributing factors and test if other p-tau epitopes are more sensitive to intermediate ADNC and thus to αSyn+AD.

Despite the high proportion of intermediate ADNC in αSyn+AD, our findings provide strong evidence that plasma GFAP is sensitive to AD-copathology in LBSD. A robust astrocytic response to β-amyloid plaques^45,46^ may in part explain the strong links found between plasma GFAP and β-amyloid^16,47^. In support, we find that plasma GFAP is sensitive to pathological alterations due to concomitant AD, and is most strongly associated with postmortem β-amyloid accumulation. Associations between GFAP and β-amyloid remained robust even after covarying for colinear tau and gliosis burden. These findings are in accordance with previous work showing plasma GFAP is associated with PET-Aβ and CSF Aβ_42_/Aβ_40_^16,17^. In LBSD, β-amyloid burden is often high and may have a synergic relationship with αSyn pathology^48^, which might in part explain the superior performance of plasma GFAP in this study compared with plasma p-tau_181_. Our analyses also point to the clinical relevance of elevated plasma GFAP, which was associated with impaired cognition (MMSE). While this study focuses on end stage disease in LBSD, it will be critical for future studies to explore whether our findings generalize to other neurodegenerative disease and other stages of disease: whether plasma GFAP is more sensitive to intermediate ADNC than p-tau_181_ in primary AD cases, and whether plasma GFAP is elevated in early/prodromal stages of disease in AD and LBSD. Likewise, it will be important for future studies to test how GFAP changes over disease course and if it predicts future cognitive decline in LBSD.

There are several caveats to consider when interpreting our findings. First, while our focus on LBSD and AD co-pathology is a strength of this study, it must be noted that pathological associations with plasma biomarkers observed here may not generalize to other conditions, such as primary AD. We also acknowledge that, despite strong associations of plasma GFAP with β-amyloid, we do not find a plasma biomarker strategy that robustly identifies concomitant ADNC in LBSD: the best ROC AUC was 0.71 using plasma GFAP. Future studies should test if plasma GFAP has added value when combined with other biomarker modalities, like CSF or PET. Second, this study focused on end stage disease, and tested how plasma levels closest to death associated with postmortem pathological accumulations. Because of this, some subjects had a substantial interval between plasma collection and death. To help account for this, models included plasma-to-death interval as a covariate and subanalyses confirmed results after excluding individuals with an interval > 5 years. Still, it will be important for future studies to track longitudinal changes in plasma biomarkers within LBSD and to test plasma GFAP and p-tau_181_ in the context of early stage LBSD. Third, plasma concentrations in LBSD may be confounded by other factors not explored in this study, including body mass index (*i.e*., blood volume) and creatinine (*i.e*., kidney functioning)^49^.

Fourth, we measured plasma p-tau_181_ concentrations using an established platform that shows excellent performance in AD^31,50^. However, future studies should test if other isoforms of plasma p-tau (217 or 231) or measures from different platforms might perform differently in LBSD. Finally, because this is a mixed pathology study, we examined the possible influence of other pathologies, but found no evidence that plasma analytes were associated with TDP-43 or CAA. However, only 3 LBSD autopsy patients had significant cerebrovascular disease, and we were not able to assess how GFAP and p-tau_181_ levels would be altered by vascular disease.

While many studies have tested AD plasma biomarkers p-tau_181_ and GFAP in the context of clinical AD, findings have not been validated in LBSD with autopsy-confirmed αSyn and AD neuropathological diagnoses. This autopsy study focuses on LBSD to evaluate AD plasma biomarkers for detecting concomitant ADNC in αSyn cases. Analyses demonstrated that plasma GFAP was sensitive to concomitant ADNC in LBSD: plasma GFAP was higher in αSyn+AD than αSyn, was sensitive to brain β-amyloid in LBSD, and was associated with global cognition in LBSD. Together, our findings demonstrate that plasma GFAP is associated with β-amyloid and concomitant AD in LBSD.

## Data Availability

All data produced in the present study are available upon reasonable request by a qualified investigator.

## Acknowledgements/Conflicts/Funding Sources

## Acknowledgments

We would like to thank the patients and families for contributing to our research and for participating in the brain donation program. We would also like to thank and acknowledge the contributions of our late colleague, John Q. Trojanowski, without whom this work would not be possible.

## Funding

This work is supported by funding from the National Institute of Aging (P01-AG066597, U19-AG062418, P30-AG072979, R01-AG054519), the National Institute of Neurological Disorders and Stroke (K23-NS114167), the Alzheimer’s Association (AARF-D-619473, AARF-D-619473-RAPID), and the Penn Institute on Aging.

## Data Sharing

Anonymized data will be shared by a reasonable request from any qualified investigator.

## Data Access

KAQC and TFT had full access to all the data in the study and take responsibility for the integrity of the data and the accuracy of the data analysis.

## Author Contributions

KC and TT contributed to the conception and design of the study. KC, DI, AC, SA, LS, EL, DW, DW, MS, AD, MG, TT all contributed to the methodology and design of the study. KC, DI, AC, SA, LS, EL, DW, DW, MS, AD, MG, TT contributed to the acquisition and analysis of data. KC and TT contributed to drafting the text or preparing the figures.

## Competing interests

Authors report no conflicts of interest relevant to this study.

## Notes

### Competing Interest Statement

The authors have declared no competing interest.

### Author Declarations

The University of Pennsylvania IRB gave ethical approval for this work.

## References

1. Irwin DJ, Lleó A, Xie SX, et al. Ante mortem cerebrospinal fluid tau levels correlate with postmortem tau pathology in frontotemporal lobar degeneration. Annals of neurology 2017;82(2):247–258.

2. Jellinger KA, Seppi K, Wenning GK, Poewe W. Impact of coexistent Alzheimer pathology on the natural history of Parkinson’s disease [Internet]. Journal of Neural Transmission 2002;109(3):329–339.Available from: https://doi.org/10.1007/s007020200027

3. Jellinger KA, Korczyn AD. Are dementia with Lewy bodies and Parkinson’s disease dementia the same disease? BMC medicine 2018;16(1):1–16.

4. Howard E, Irwin DJ, Rascovsky K, et al. Cognitive Profile and Markers of Alzheimer Disease–Type Pathology in Patients With Lewy Body Dementias [Internet]. Neurology 2021;96(14):e1855 LP–e1864.Available from: http://n.neurology.org/content/96/14/e1855.abstract

5. Shellikeri S, Cho S, Cousins KAQ, et al. Natural speech markers of Alzheimer’s disease co-pathology in Lewy body dementias [Internet]. Parkinsonism & Related Disorders 2022;Available from: https://www.sciencedirect.com/science/article/pii/S1353802022002486

6. Lemstra AW, Beer MH de, Teunissen CE, et al. Concomitant AD pathology affects clinical manifestation and survival in dementia with Lewy bodies [Internet]. Journal of Neurology, Neurosurgery & Psychiatry 2017;88(2):113 LP–118.Available from: http://jnnp.bmj.com/content/88/2/113.abstract

7. Walker IM, Cousins KA, Siderowf A, et al. Non-tremor motor dysfunction in Lewy body dementias is associated with AD biomarkers [Internet]. Parkinsonism & Related Disorders 2022;100:33–36.Available from: https://www.sciencedirect.com/science/article/pii/S1353802022001699

8. Irwin DJ, Xie SX, Coughlin D, et al. CSF tau and, 8-amyloid predict cerebral synucleinopathy in autopsied Lewy body disorders. Neurology 2018;90(12):e1038–e1046.

9. Tropea TF, Chen-Plotkin A. Are Parkinson’s Disease Patients the Ideal Preclinical Population for Alzheimer’s Disease Therapeutics? Journal of Personalized Medicine 2021;11(9):834.

10. Cousins KAQ, Arezoumandan S, Shellikeri S, et al. CSF Biomarkers of Alzheimer Disease in Patients With Concomitant a-Synuclein Pathology [Internet]. Neurology 2022;10.1212/WNL.0000000000201202.Available from: http://n.neurology.org/content/early/2022/08/30/WNL.0000000000201202.abstract

11. Irwin DJ, Fedler J, Coffey CS, et al. Evolution of Alzheimer’s Disease Cerebrospinal Fluid Biomarkers in Early Parkinson’s Disease. Annals of Neurology 2020;88(3):574–587.

12. Brickman AM, Manly JJ, Honig LS, et al. Plasma p-tau181, p-tau217, and other blood-based Alzheimer’s disease biomarkers in a multi-ethnic, community study. Alzheimer’s and Dementia 2021;17(8):1–12.

13. Thijssen EH, La Joie R, Strom A, et al. Plasma phosphorylated tau 217 and phosphorylated tau 181 as biomarkers in Alzheimer’s disease and frontotemporal lobar degeneration: a retrospective diagnostic performance study. The Lancet Neurology 2021;20(9):739–752.

14. Ishiki A, Kamada M, Kawamura Y, et al. Glial fibrillar acidic protein in the cerebrospinal fluid of Alzheimer’s disease, dementia with Lewy bodies, and frontotemporal lobar degeneration. Journal of neurochemistry 2016;136(2):258–261.

15. Heneka MT, Carson MJ, El Khoury J, et al. Neuroinflammation in Alzheimer’s disease. The Lancet Neurology 2015;14(4):388–405.

16. Pereira JB, Janelidze S, Smith R, et al. Plasma GFAP is an early marker of amyloid-,8 but not tau pathology in Alzheimer’s disease. Brain 2021;

17. Cicognola C, Janelidze S, Hertze J, et al. Plasma glial fibrillary acidic protein detects Alzheimer pathology and predicts future conversion to Alzheimer dementia in patients with mild cognitive impairment. Alzheimer’s Research & Therapy 2021;13(1):1–9.

18. Chouliaras L, Thomas A, Malpetti M, et al. Differential levels of plasma biomarkers of neurodegeneration in Lewy body dementia, Alzheimer’s disease, frontotemporal dementia and progressive supranuclear palsy [Internet]. Journal of Neurology, Neurosurgery & Psychiatry 2022;93(6):651–658.Available from: http://jnnp.bmj.com/content/93/6/651.abstract

19. Hall S, Janelidze S, Zetterberg H, et al. Cerebrospinal fluid levels of neurogranin in Parkinsonian disorders. Movement Disorders 2020;35(3):513–518.

20. Gonzalez MC, Ashton NJ, Gomes BF, et al. Association of Plasma p-tau181 and p-tau231 Concentrations With Cognitive Decline in Patients With Probable Dementia With Lewy Bodies [Internet]. JAMA Neurology 2022;79(1):32–37.Available from: https://doi.org/10.1001/jamaneurol.2021.4222

21. Toledo JB, Van Deerlin VM, Lee EB, et al. A platform for discovery: the University of Pennsylvania integrated neurodegenerative disease biobank. Alzheimer’s & dementia 2014;10(4):477–484.

22. Stuart EA, King G, Imai K, Ho D. MatchIt: nonparametric preprocessing for parametric causal inference [Internet]. Journal of Statistical Software 2011;42(8):1–28.Available from: http://www.jstatsoft.org/v42/i08/

23. Lee EB. Integrated neurodegenerative disease autopsy diagnosis. Acta neuropathologica 2018;135(4):643–646.

24. Montine TJ, Phelps CH, Beach TG, et al. National institute on aging-Alzheimer’s association guidelines for the neuropathologic assessment of Alzheimer’s disease: A practical approach. Acta Neuropathologica 2012;123(1):1–11.

25. Thal DR, Rüb U, Orantes M, Braak H. Phases of A,8-deposition in the human brain and its relevance for the development of AD. Neurology 2002;58(12):1791–1800.

26. Braak H, Braak E. Neuropathological stageing of Alzheimer-related changes. Acta neuropathologica 1991;82(4):239–259.

27. Mirra SS, Heyman A, McKeel D, et al. The Consortium to Establish a Registry for Alzheimer’s Disease (CERAD): Part II. Standardization of the neuropathologic assessment of Alzheimer’s disease. Neurology 1991;41(4):479.

28. McKeith IG, Boeve BF, Dickson DW, et al. Diagnosis and management of dementia with Lewy bodies: Fourth consensus report of the DLB Consortium. Neurology 2017;89(1):88–100.

29. Skrobot OA, Attems J, Esiri M, et al. Vascular cognitive impairment neuropathology guidelines (VCING): the contribution of cerebrovascular pathology to cognitive impairment. Brain 2016;139(11):2957–2969.

30. Mackenzie IRA, Neumann M, Baborie A, et al. A harmonized classification system for FTLD-TDP pathology. Acta neuropathologica 2011;122(1):111–113.

31. Tropea TF, Waligorska T, Xie SX, et al. Plasma Phosphorylated Tau181 is a Biomarker of Alzheimer’s Disease Pathology and Associated with Cognitive and Functional Decline [Internet]. Annals of clinical and translational neurology 2022;Available from: http://dx.doi.org/10.2139/ssrn.4007185

32. Cousins KAQ, Shaw LM, Chen-Plotkin A, et al. Distinguishing Frontotemporal Lobar Degeneration Tau From TDP-43 Using Plasma Biomarkers [Internet]. JAMA Neurology 2022;79(11):1155–1164.Available from: https://doi.org/10.1001/jamaneurol.2022.3265

33. Chatterjee P, Pedrini S, Stoops E, et al. Plasma glial fibrillary acidic protein is elevated in cognitively normal older adults at risk of Alzheimer’s disease. Translational psychiatry 2021;11(1):1–10.

34. Ben-Shachar M, Lüdecke D, Makowski D. effectsize: Estimation of Effect Size Indices and Standardized Parameters [Internet]. Journal of Open Source Software 2020;5(56):2815.Available from: https://github.com/easystats/effectsize

35. Olejnik S, Algina J. Generalized eta and omega squared statistics: measures of effect size for some common research designs. Psychological methods 2003;8(4):434.

36. Thiele C, Hirschfeld G. Cutpointr: Improved estimation and validation of optimal cutpoints in R. Journal of Statistical Software 2021;98(11):1–27.

37. Lantero Rodriguez J, Karikari TK, Suárez-Calvet M, et al. Plasma p-tau181 accurately predicts Alzheimer’s disease pathology at least 8 years prior to post-mortem and improves the clinical characterisation of cognitive decline [Internet]. Acta Neuropathologica 2020;140(3):267–278.Available from: https://doi.org/10.1007/s00401-020-02195-x

38. Weinshel S, Irwin DJ, Zhang P, et al. Appropriateness of Applying Cerebrospinal Fluid Biomarker Cutoffs from Alzheimer’s Disease to Parkinson’s Disease. Journal of Parkinson’s Disease 2022;12:1155–1167.

39. Cousins KAQ, Shaw LM, Shellikeri S, et al. Elevated plasma phosphorylated tau 181 in amyotrophic lateral sclerosis. Annals of Neurology 2022;92(5):807–818.

40. Benedet AL, Milà-Alomà M, Vrillon A, et al. Differences between plasma and cerebrospinal fluid glial fibrillary acidic protein levels across the Alzheimer disease continuum. JAMA neurology 2021;78(12):1471–1483.

41. Ashton NJ, Janelidze S, Al Khleifat A, et al. A multicentre validation study of the diagnostic value of plasma neurofilament light. Nature communications 2021;12(1):1–12.

42. Smirnov DS, Ashton NJ, Blennow K, et al. Plasma biomarkers for Alzheimer’s Disease in relation to neuropathology and cognitive change. Acta neuropathologica 2022;143(4):487–503.

43. Thijssen EH, La Joie R, Wolf A, et al. Diagnostic value of plasma phosphorylated tau181 in Alzheimer’s disease and frontotemporal lobar degeneration. Nature Medicine 2020;26(3):387–397.

44. Karikari TK, Emeršič A, Vrillon A, et al. Head-to-head comparison of clinical performance of CSF phospho-tau T181 and T217 biomarkers for Alzheimer’s disease diagnosis. Alzheimer’s & Dementia 2021;17(5):755–767.

45. Kamphuis W, Middeldorp J, Kooijman L, et al. Glial fibrillary acidic protein isoform expression in plaque related astrogliosis in Alzheimer’s disease [Internet]. Neurobiology of Aging 2014;35(3):492–510.Available from: https://www.sciencedirect.com/science/article/pii/S0197458013004375

46. Nagele RG, Wegiel J, Venkataraman V, et al. Contribution of glial cells to the development of amyloid plaques in Alzheimer’s disease [Internet]. Neurobiology of Aging 2004;25(5):663–674.Available from: https://www.sciencedirect.com/science/article/pii/S0197458004001034

47. De Bastiani MA, Bellaver B, Brum W, et al. Hippocampal GFAP-positive astrocyte responses to amyloid and tau pathologies [Internet]. 2022;Available from: http://europepmc.org/abstract/PPR/PPR462653 https://doi.org/10.1101/2022.02.25.481812

48. Lashley T, Holton JL, Gray E, et al. Cortical a-synuclein load is associated with amyloid-,8 plaque burden in a subset of Parkinson’s disease patients. Acta neuropathologica 2008;115(4):417–425.

49. Pichet Binette A, Janelidze S, Cullen N, et al. Confounding factors of Alzheimer’s disease plasma biomarkers and their impact on clinical performance [Internet]. Alzheimer’s & Dementia 2022;Available from: https://alz-journals.onlinelibrary.wiley.com/doi/10.1002/alz.12787,

50. Karikari TK, Pascoal TA, Ashton NJ, et al. Blood phosphorylated tau 181 as a biomarker for Alzheimer’s disease: a diagnostic performance and prediction modelling study using data from four prospective cohorts. The Lancet Neurology 2020;19(5):422–433.

